# HbA1c-based diagnosis of type 2 diabetes and complication risk are distorted in British south Asians due to HbE thalassaemia trait

**DOI:** 10.64898/2026.03.25.26348217

**Authors:** Sam Hodgson, Veline L’Esperance, Miriam Samuel, Moneeza Siddiqui, Daniel Stow, Camila Armirola-Ricuarte, Genes & Health Research Team, David A. van Heel, Rohini Mathur, TJ Mckinley, Inês Barroso, Joseph Taylor, Sarah Finer

**Affiliations:** Wolfson Institute of Population Health, Queen Mary University of London; (1) Department of Clinical and Biomedical Sciences, Faculty of Health and Life Sciences, University of Exeter Medical School, University of Exeter. (2) Exeter Centre of Excellence for Diabetes Research (EXCEED), University of Exeter Medical School; Blizard Institute, Queen Mary University of London; Precision Healthcare University Research Institute (PHURI), Queen Mary University of London

## Abstract

**Background:** Genetic variants impacting red blood cell biology disrupt the relationship between glycaemia and glycated haemoglobin (HbA1c), with implications for diagnosis and management of type 2 diabetes (T2D). Thalassaemia trait is estimated to affect 350 million people globally, but its impact on T2D and related outcomes is not clear.

**Methods:** We explored associations between thalassaemia trait, HbA1c, and T2D diagnosis and complications in 43,088 British Bangladeshi and Pakistani participants in the Genes & Health study with linked multisource England National Health Service (NHS) electronic health record data and whole exome sequencing.

**Findings:** 2,490 participants (5.8%) were heterozygous carriers of ClinVar pathogenic / likely pathogenic thalassaemia variants, however 3 in 4 of these were not diagnosed with thalassaemia in their NHS health records. rs33950507, a common variant causal for *HbE* thalassaemia, was associated with increased HbA1c (beta=0.13, 95%CI:0.08-0.18, p=7.8×10^−8^), but not glucose levels (beta=0.01, 95%CI:-0.04-0.06, P=0.72). rs33950507 was associated with increased hazards of prediabetes (HR=1.38, 95%CI:1.26-1.52, p=2.2×10^−6^) and T2D (HR=1.11, 95%CI:1.01-1.22, p=0.03), and reduced hazards of diabetic eye disease (HR=0.74, 95%CI:0.56-0.96, p=0.02) and cerebrovascular disease (HR=0.44, 95%CI:0.20-0.94, p=0.03). Sensitivity analyses suggested mediation by overdiagnosis and overtreatment of T2D.

**Interpretation:** Alternatives to HbA1c, and/or precision medicine approaches to defining and managing pre-diabetes and T2D. These findings have global impact, and are particularly relevant to individuals from ancestral groups among whom erythrocytic traits are more common but often undiagnosed.

**Funding:** Wellcome Trust, MRC, NIHR, Barts Charity, Genes & Health Industry Consortium

**RESEARCH IN CONTEXT:** *Evidence before this study:* The role of genetic variants that disrupt red blood cell biology in distorting the relationship between glycaemia and glycated haemoglobin (HbA1c) is increasingly recognised. Recently, variants in *G6PD* and *PIEZO1,* associated with lower HbA1c via red blood cell mechanisms independent of blood glucose, have been shown to associate with increased progression of diabetes complications. The clinical consequences of HbA1C disruption from thalassaemia trait remain poorly understood, despite thalassaemia trait affecting 350 million individuals worldwide, and being known to interfere with HbA1c measurement. We searched PUBMED and EMBASE for “thalass* & (diabet* | “hba1” | glycat*)” from inception to December 2025, identifying 984 studies. While several studies described distortion of HbA1c relative to blood glucose in thalassaemia trait, we identified none which reported on the clinical consequences, or on the diagnosis of diabetes or its complications.

*Added value of this study:* Thalassemia affects over 350 million people globally. We defined thalassaemia trait as individuals heterozygous for ClinVar pathogenic / likely pathogenic variants in whole exome sequence data in the Genes & Health study of British Pakistani and Bangladeshi individuals (n = 2,490 / 43,098). We highlight that only 1 in 4 of these genetically-defined thalassaemia trait carriers had a clinical diagnosis in their NHS health records, suggesting that thalassaemia is under-recognised in clinical practice in this population. A clinical prediction tool using routine. blood tests (ThalPred), was also only able to identify only one quarter of thalassaemia trait carriers. *HbE* thalassaemia trait was associated with increased HbA1c, contrasting to findings previously reported for *G6PD* and PIEZO1 which show opposite directionality. Thalassemia trait was also associated with increased hazards of prediabetes and type 2 diabetes (T2D) diagnosis, but was not associated with changes in serum glucose. *HbE* trait was associated with reduced hazards of complications including diabetic eye and cerebrovascular disease, but these associations were lost after adjustment for HbA1c, T2D diagnosis, and antidiabetic drug exposure. These findings suggest that the distortion of HbA1c leads to over-diagnosis and over-treatment, and this mediates the association with complications.

*Implications of all the available evidence:* This study adds further evidence to a growing literature emphasising the distortion between HbA1c and glycaemia in genetic conditions affecting red cell biology. Our findings suggest there is need for alternatives to HbA1c, and/or precision medicine approaches, to diagnose and manage prediabetes and type 2 diabetes accurately and equitably. These findings are particularly relevant in Asian ancestral groups, among whom poorer diabetes outcomes and red cell traits such as thalassaemia are common.

## Introduction

The majority of modern type 2 diabetes care rests on the assumption that HbA1c accurately reflects glycaemia, yet inherited variation in red blood cell biology may systematically distort this relationship in millions of individuals worldwide^1,2^. Type 2 diabetes disproportionately affects individuals of south Asian ancestry^3,4^, among whom haemoglobin variants shaped by historical malaria selection are common^5–7^. Across health systems, HbA1c underpins diagnosis, risk stratification and treatment intensification as a metric of glycaemia^8–11^ .

Erythrocytic traits affecting red blood cell biology are common: an estimated 350 million people worldwide carry thalassemia trait,^1,2^ for example. While many type 2 diabetes treatment guidelines suggest avoiding use of HbA1c in the context of erythrocytic conditions, and, in trait form, these are often clinically silent and undiagnosed^1,2,12^ and there is little clarity on what alternative tests should be used^10^. Recent evidence shows HbA1c-driven diagnosis and management of type 2 diabetes may impact outcomes from the condition, as these individuals often have lower HbA1c levels for a given blood glucose, as has been demonstrated in G6PD deficiency, alpha-thalassaemia, and mutations in the PIEZO1 gene^1,13–15^. These erythrocytic traits have also been associated with reduced rates of type 2 diabetes diagnosis^1,2^, and increased risk of diabetes-related complications such as eye disease^1^, plausibly due to delayed diagnosis and under-treatment arising from underestimation of glycaemia by monitoring predicated on HbA1c values not reflective of an individual’s true glycaemic state. Furthermore, because these erythrocytic traits are more common in ancestral groups who tend to have poorer outcomes in type 2 diabetes^16–18^, such as African and south Asian ancestry individuals, there is a risk that inaccuracy in HbA1c from red cell traits could contribute to or worsen health inequalities.

Conversely, disruption to protein structure in thalassaemia traits such as HbE, a specific form of beta-thalassaemia common in south Asia (prevalence ∼5%) and south-east Asia (prevalence 10-30%)^19^, may be associated with increased HbA1c readings relative to blood glucose, likely due to changes in cationic charge associated with protein structure alteration which interfere with high performance liquid chromatography (HPLC) assays for HbA1c^20,21^.

The aim of this study is therefore to systematically explore the association of genetically-ascertained HbE thalassaemia trait with HbA1c levels, progression to T2D diagnosis, and diabetes-related complications, in a large population-based, longitudinal cohort study of British Bangladeshi and Pakistani individuals in the UK.

## Methods

### Study population

Genes & Health is a long-term population cohort study of British Bangladeshi and British Pakistani individuals aged >16 years, linking longitudinal electronic health record data from primary and secondary NHS healthcare providers with high quality genetic data (both imputed genotype and whole exome sequencing (WES)) and multi-omic data^22,23^. As of March 2026 there were ∼75,000 participants recruited.

### Cohort selection

In this study, we included all participants aged 16 years or over at the time of recruitment with whole exome sequence data, linked primary and secondary care NHS data (with records extending from birth to most recent data extraction), and at least one recorded laboratory full blood count. We excluded individuals with diagnostic clinical codes of sickle cell disease, who had ever received one (or more) blood transfusion(s), who were homozygous or compound heterozygous carriers for pathogenic thalassaemia variants (see variant definitions below), or who had evidence in the electronic health record of type 1 diabetes, monogenic diabetes, or secondary causes of diabetes. Further details of numbers of participants included and excluded are given in **Fig 1**.

**Figure 1:**
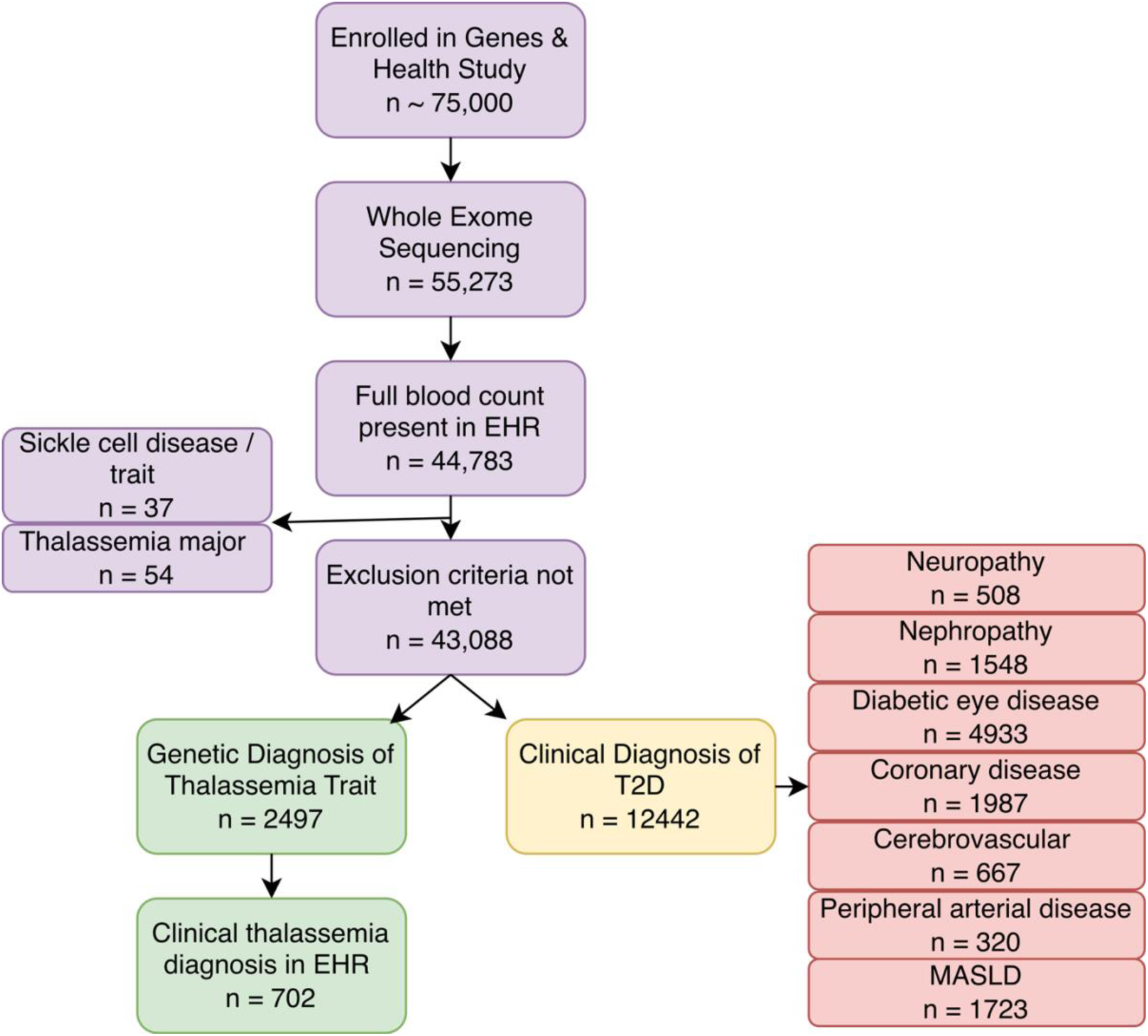
Participant flow diagram of included individuals with whole exome sequence data (purple), clinical and genetic diagnosis of thalassaemia trait (green), and clinical diagnosis of type 2 diabetes and its complications (red) EHR: electronic health record; T2D: type 2 diabetes; MASLD: metabolic dysfunction-associated steatotic liver disease.

### Whole exome sequencing

We used whole exome sequence data, currently available for 55,273 volunteers, for genetic analysis. Exome sequencing steps, quality control, and variant annotation have been described in detail elsewhere^24^, and further details are given in **the Supplementary methods.**

### ClinVar variant definitions

We defined causal thalassaemia variants using ClinVar, including variants with a label of “Thalassaemia” which were defined as either “Pathogenic” or “Likely pathogenic”. We excluded variants which included a label of “sickle cell” or “Sickle cell anaemia”. We identified heterozygous, homozygous and compound heterozygous individuals for these variants in whole exome sequence data using Plink V2.0^25^. Because we identified only 16 individuals with mutations in *HBA1/HBA2* (ie, alpha-thalassaemia causing variants), as would be expected based on global disease epidemiology, we restricted analyses to beta-thalassaemia mutations (ie, those in *HBB)(***Table S1-2**).

### EHR case ascertainment

We defined a clinical diagnosis of thalassaemia from the primary or secondary care NHS data as an ever-recorded ICD10 code or SNOMED code relating to thalassaemia or related traits **(Table S3).** Type 2 diabetes, prediabetes, and related complications (**Supplementary text)** were similarly defined according to the clinical codes in **Table S3;** only codes first appearing above 18 years of age were included. Codes used to exclude individuals with sickle cell anaemia, type 1 diabetes, MODY diabetes, or secondary causes of diabetes are given in **Table S3.**

### EHR clinically measured quantitative trait definitions

We obtained laboratory test values from routinely collected electronic health record data, obtained from primary or secondary care. This longitudinal quantitative data, which includes repeated measures where clinically performed, was curated as described previously^2^. In brief, outliers 10 standard deviations above or below the mean were excluded, and individual-level lifetime median values used for analysis.

Because glycaemia is impacted by antidiabetic treatment, for HbA1c and glucose values, we restricted analyses to values obtained before the first date of T2D diagnosis or initiation of antidiabetic treatment (**Supplementary text, Table S4)**. We additionally excluded any values within a 9-month window before or after pregnancy-related codes in the health record data, because of changing patterns in glycaemia and red blood cell turnover during pregnancy. Further details on HbA1c assays used are given in the **supplementary text.**

### ThalPred prediction of thalassaemia trait

Given thalassemia screening is not routine in the NHS and underdiagnosis is common^26^ we explored whether ThalPred^27^, a published thalassaemia risk prediction tool which uses laboratory values of haemoglobin (Hb), mean cell volume (MCV), mean corpuscular haemoglobin concentration (MCHC), red blood cell (RBC) count and red cell distribution width (RDW) could aid in clinical ascertainment or discrimination of genetically-determined thalassaemia in individuals with a microcytic, hypochromic anaemia (n = 3125). We used individual-level lifetime median values for each algorithmic input.

### Association testing

We defined “common” genetic variants as those with a minor allele frequency (MAF) >1%, and “rare” variants as those with a MAF < 1%. Because we had excluded homozygous individuals, our statistical analyses assess binarized associations between heterozygotes (thalassaemia trait / carriers) and non-carriers. Details of common variant and rare variant multivariable association testing using REGENIE are provided in the **supplementary text.**

### Meta-analysis

Where possible, for HbA1c analyses, we externally validated results and meta-analysed with publicly available summary statistics from other studies^28,29^ using fixed-effect models: further details are provided in the **supplementary text**.

### Sensitivity analyses

To explore whether associations between *HbE* genotype and T2D complications were influenced by HbA1c, T2D diagnosis, and treatment, we performed a series of sensitivity analyses following a mediation analysis framework^30^. We first defined plausible mechanisms using a direct acyclic graph (DAG). We then performed stepwise sensitivity analyses, adjusting main analyses (survival models for the relationship between T2D complications ∼ *HbE* genotype) for, in turn, HbA1c, T2D diagnosis state (ever/never), and exposure to 6 or more oral antidiabetic drugs (ever/never). We interpreted attenuation of the previously statistically significant signal as weak evidence that each additional covariate, in turn, may be mediating the association between HbE trait and outcomes. We compared these outcomes to a common but unrelated trait, acne vulgaris, as a negative control. Details of further sensitivity analyses are presented in the **Supplementary Text.**

## Results

### Thalassaemia trait common in British south Asians

In total, 43,088 British Bangladeshi and British Pakistani participants were eligible for inclusion in our analyses **(Fig 1**, **Table 1).**

**Table 1:** Demographic characteristics of included participants.

Of 274 genetic variants classified as “Pathogenic” or “Likely pathogenic” for thalassaemia on ClinVar, 33 were present in whole exome sequence data in our analysis (**Table S1).** In addition to these variants, we identified five further ultra-low-frequency, high-confidence predicted loss-of-function variants present in Genes & Health which have not been described on Clinvar (three in *HBB*, one in *HBA1*, and one in *HBA2*)**(Table S2)**. Because only 16 individuals had mutations in *HBA1*/*HBA2* across all included participants, we focused analyses on variants in *HBB*, including both ClinVar and high-confidence loss-of-function variants in analyses.

### Pathogenic thalassaemia variants are associated with hypochromic, microcytic anaemia

We explored the association of Clinvar Pathogenic / Likely pathogenic variants with red blood cell traits, to confirm these were associated with a clinical picture of thalassaemia trait.

For a single common variant with a minor allele frequency (MAF) > 1% (*rs33950507*, causal for *HbE* trait) we performed association testing using multivariable regression models adjusted for age, age^2^, sex, ancestry, and the first 20 genetic principal components. For rare variants with MAF <1%, we collapsed variants into an aggregated “mask” and explored the association of these aggregates with phenotypic traits using models adjusted for the same covariates as the single-variant analyses. As would be expected, pathogenic variants for thalassaemia across the allele frequency spectrum were associated with a microcytic, hypochromic anaemia, increased red cell count, changes in HbA, HbA2 and HbF percentages, and features of haemolysis **(Fig S1A).** Among 606 participants with recorded HbE percentages in the EHR, 590 rs33950507 heterozygotes and 15 homozygotes had HbE percentages in the expected elevated ranges **(Fig S2),** suggesting rs33950507 is associated with the expected HbE thalassaemia phenotype.

### Thalassemia trait is underdiagnosed in British south Asians

In total, we identified n=2490 (5.2%) individuals heterozygous for ClinVar Pathogenic/Likely Pathogenic variants and hence genetically determined thalassaemia trait carriers. Of these individuals, only 702 (28.1%) had a clinical diagnosis relating to thalassaemia in the electronic health record data from either primary or secondary care. Therefore, overall, 71.9% of individuals with thalassaemia trait did not have an associated diagnosis in their electronic health care record data.

### A validated clinical prediction tool did not aid the clinical ascertainment of thalassemia trait

We used an established thalassaemia prediction algorithm, ThalPred^27^, to explore whether a clinical risk prediction tool might improve ascertainment of clinically defined EHR cases, given the underdiagnosis in health records of trait carriers. This algorithm attempts to discriminate thalassaemia trait from iron deficiency anaemia using laboratory values for Hb, RBC count, MCV, MCHC, and RDW in individuals with microcytic anaemia. Among a subset of 3125 individuals with microcytic anaemia, 1020 had genetically-determined thalassaemia trait. However, the algorithm also identified an additional 1764 individuals as likely thalassaemia trait, but who did not have genetic causal variants, i.e. false positives (specificity = 22.1%; positive predictive value = 35.5%). Furthermore, as the majority of individuals with thalassaemia trait did not have a microcytic anaemia to enable use of ThalPred, overall only 34.3% of individuals with a genetic diagnosis of thalassaemia trait were identified using this method.

### Genetic variants which cause thalassaemia are associated with changes in HbA1c but not glucose

We next examined the association of common and rare ClinVar-defined thalassaemia variants with glycaemic traits and type 2 diabetes states. In addition to associations with red cell traits (**Fig S1)**, rs33950507 was associated with increased lifetime median HbA1c (beta = 0.13, 95% CI = 0.08 – 0.18, p = 7.8 × 10^−8^), but was not associated with changes in non-fasting glucose (beta = 0.01, 95% CI = -0.04 – 0.06, p = 0.72) **(Fig 2,Table S5),** suggesting the associations with HbA1c may be independent of glycaemic state. When we attempted to replicate this finding in UK Biobank, we discovered that research-based HPLC-measured baseline HbA1c for individuals with HbE trait were excluded from their protocols with an “analyser result deemed not reportable” flag. The association of rs33950507 with HbA1c could be replicated using publicly available summary statistics from 2872 south Asians from four cohort studies compiled as part of Flannick et al’s exome-wide association study^28^ (beta = 0.05, 95% CI = 0.002 – 0.10, p = 0.01). The association between rs33950507 and HbA1c was directionally consistent and conventionally statistically significant (p<0.05) after meta-analysis (beta = 0.09, 95% CI = 0.06 – 0.13, p = 5.5 × 10^−7^).

**Fig 2:**
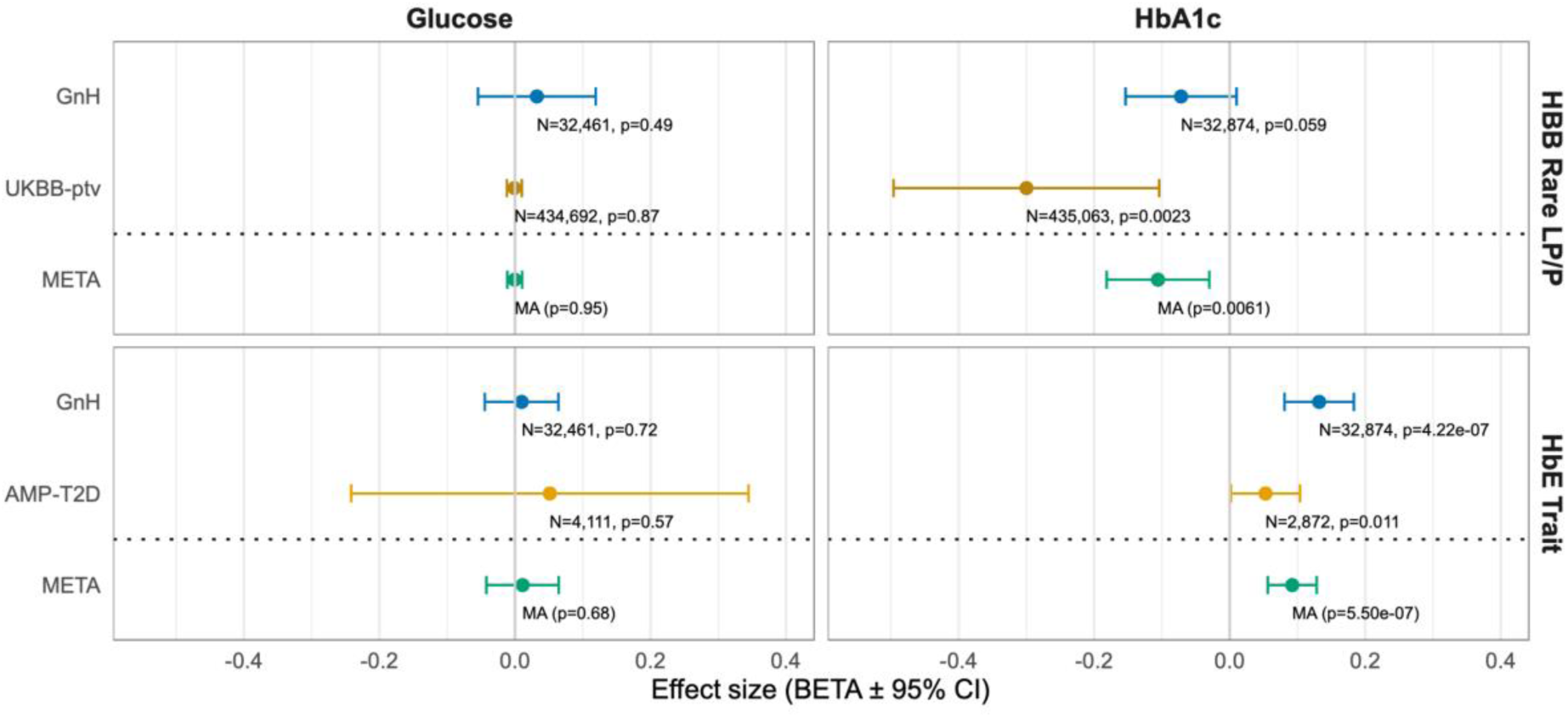
Association of genetically determined thalassaemia trait with glycaemic traits, with external replication, shows heterogeneous direction of effect on HbA1c between common and rare HBB variants, with no association with glucose. Results are meta-analysed between GnH and external available summary data using fixed-effect models. For rare Clinvar likely pathogenic / pathogenic variants in HBB with minor allele frequency < 0.01 (HBB Rare LP/P), results were meta-analysed with those described from the most closely aligned mask available on the AstraZeneca Phewas portal, comprised of UK Biobank summary statistics (UKBB-ptv: protein-truncating variants). For the single HbE trait variant (rs33950507) results were meta-analysed with whole exome sequenced results published as part of the AMP-T2D study.

In contrast, and consistent with previously reported findings for erythrocytic variants, aggregate masks of rare thalassaemia causing variants with MAF <1% were associated with a non-significant trend toward lower HbA1c (beta = -0.08, 95% CI = -0.16 – 0.003, p = 0.059) **(Fig 2),** suggesting mechanistically distinct pathways. This result could also be partially replicated using publicly available summary statistics from predicted loss of function (protein-truncating variant) rare variant masks in UK Biobank^29^ (beta = -0.30, 95% CI = -0.10 - -0.49, p = 0.002), although the variants contained within this mask do not necessarily align exactly with those used in Genes & Health mask definitions. The meta-analysed result was statistically significant (beta = -0.11, 95% CI = -0.18 - -0.03, p = 0.006).

### Genetically-determined thalassaemia trait is associated with risk of prediabetes and type 2 diabetes diagnoses

In addition to its association with increased HbA1c **(Fig 2)**, rs33950507 was associated with increased hazards of prediabetes (HR = 1.38, 95% CI = 1.26 – 1.52, p = 2.2 × 10^−6^) and type 2 diabetes (HR = 1.11, 95% CI = 1.01 – 1.22, p = 0.03) (**Fig 3)**. Both these associations were also significant in univariate survival models (illustrative Kaplan-Meier plots presented in **Fig 3)**, and in keeping with previous reports of rs33950507 being associated with increased odds of type 2 diabetes in the largest published multi-ancestry meta-analysis to date(OR = 1.44, 95% CI – 1.14 – 1.82, p = 0.003)^31^.

**Fig 3:**
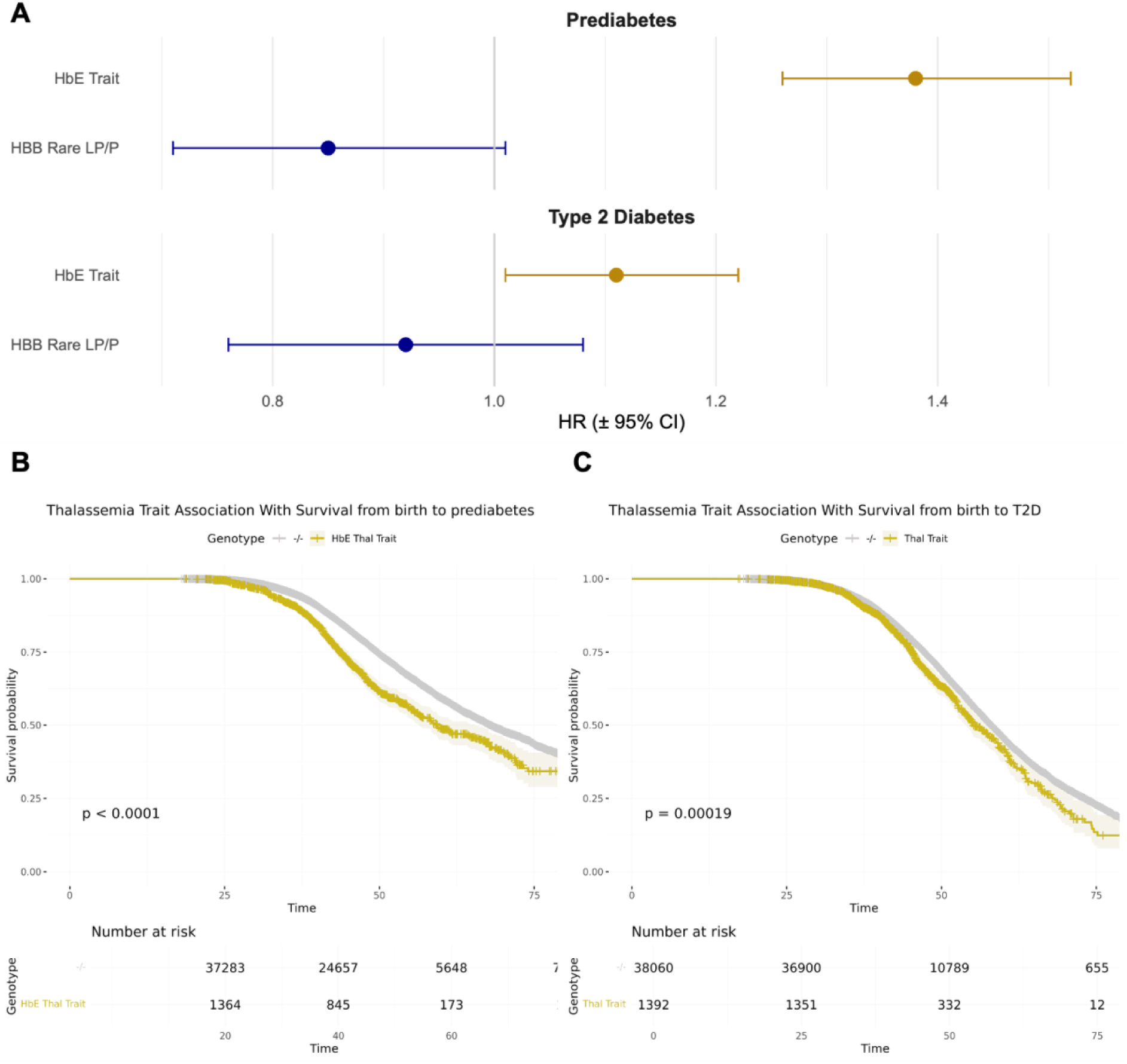
Association of genetically determined thalassaemia trait with progression from birth to prediabetes and T2D. A: forest plots depicting hazard ratio and 95% confidence intervals (HR + 95% CI) estimated from multivariable Cox proportional hazards models adjusted for age at recruitment, age at recruitment squared, sex, genetically inferred ancestry, and the first 20 genetic principal components, presented separately for rare Clinvar likely pathogenic / pathogenic variants in HBB with minor allele frequency < 0.01 (HBB Rare LP/P), and a single common variant (rs33950507, causal for HbE trait). B: Kaplan-Meier plot depicting survival from birth to date of prediabetes diagnosis, stratified by HbE (rs33950507) genotype – HbE trait carriers (blue line) vs unaffected individuals (red line), truncated at age = 75 years (∼10% of original sample remaining at risk). C: Kaplan-Meier plot depicting survival from birth to date of type 2 diabetes diagnosis, stratified by HbE genotype as in B, truncated at age = 75 years (∼10% of original sample at risk).

Collapsed masks of HBB pathological rare variants with MAF <1% were not associated with progression from birth to either prediabetes or diabetes, although a non-significant trend towards lower hazards of prediabetes was observed (HR = 0.85, 95% CI = 0.71 – 1.01, p = 0.079) (**Fig 3)**, in keeping with previous reports in the literature of the association of erythrocytic variants with delays in diagnosis of T2D^1,2^.

### HbE thalassaemia causal variants are associated with reduced progression to diabetes-related complications

Finally, we examined the association of pathogenic thalassaemia variants with progression to micro- and macrovascular complications of type 2 diabetes in survival models from birth **(Fig 4A, S3).** rs33950507 was associated with lower hazards of diabetic eye disease (HR = 0.74, 95% CI = 0.56 – 0.96, p = 0.02), cerebrovascular disease (HR = 0.44, 95% CI = 0.20 – 0.94, p = 0.03), and metabolic dysfunction-associated steatotic liver disease (MASLD) (HR = 0.52, 95% CI = 0.32 – 0.84, p = 0.004), but increased hazards of heart failure (HR = 2.01, 95% CI = 1.00 – 4.02, p = 0.048). However, none of these associations remained significant after correction for multiple testing (p < 0.0025).

**Fig 4.**
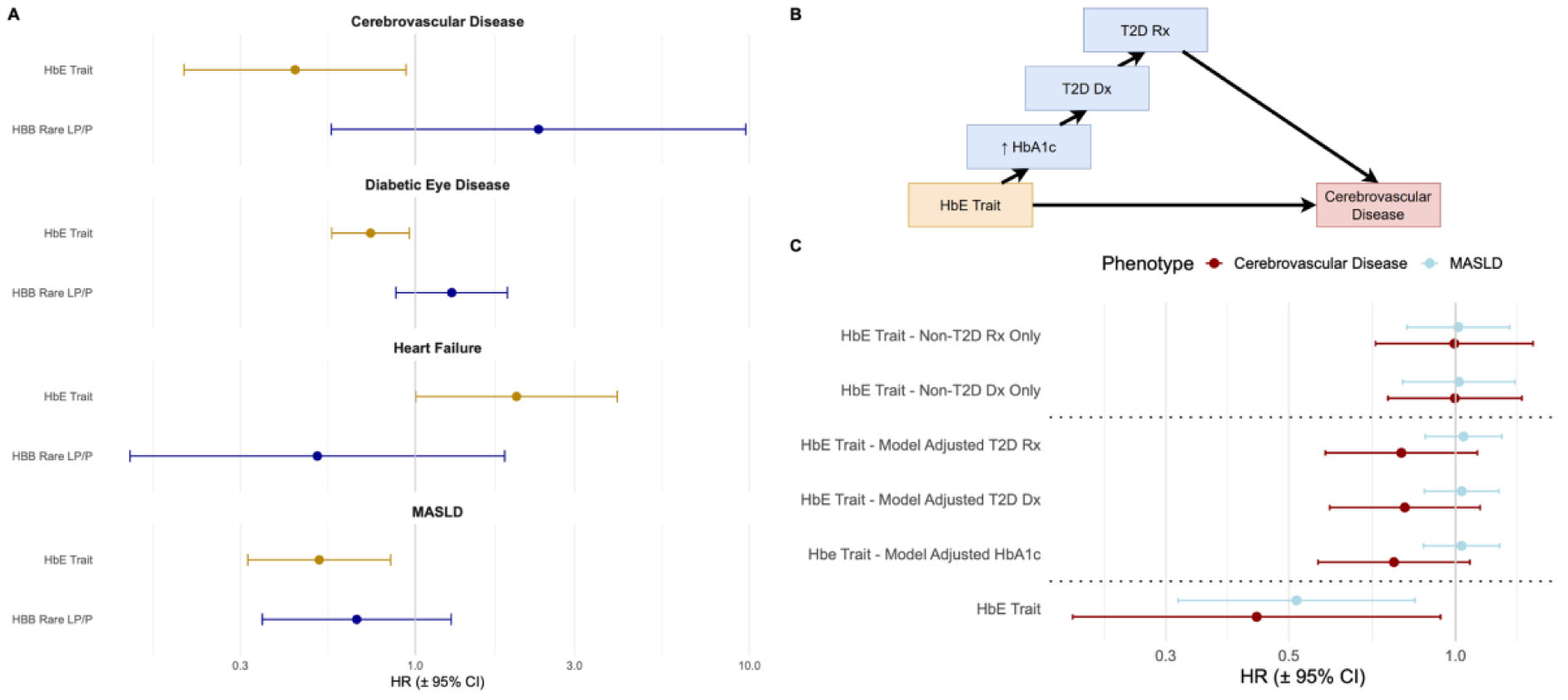
A: Association of pathogenic thalassaemia trait variants with progression from birth to selected complications. A: Results are presented as hazard ratio (HR) +- 95% confidence interval, estimated from multivariable Cox proportional hazards models adjusted for age at recruitment, age at recruitment squared, sex, genetically inferred ancestry, and the first 20 genetic principal components, presented separately for rare Clinvar likely pathogenic / pathogenic variants in HBB with minor allele frequency < 0.01 (HBB Rare LP/P), and a single common variant (rs33950507, causal for HbE trait). MASLD: metabolic dysfunction-associated steatotic liver disease. B: Schematic directed acyclic graph (DAG) used as the schema for sensitivity analyses following a mediation analytic framework. C: Results from sensitivity analyses based on the analyses in (A), additionally adjusted for HbA1c (lifetime median value), T2D diagnosis (T2D Dx), T2D treatment with antidiabetic drugs on at least 6 occasions (T2D Rx); or without additional adjustment, but restricted only to individuals free from T2D diagnosis, or free from T2D treatment.

### The association between HbE and diabetic complications may be mediated by HbA1c, T2D diagnosis, and T2D treatment

As would be expected, the negative association between rs33950507 and diabetic eye disease was attenuated and no longer significant after adjustment for T2D status. However, in a series of sensitivity analyses following a mediation analysis framework, similar attenuations were observed for both cerebrovascular disease (HR = 0.81, 95% CI = 0.59 – 1.11, p = 0.45) and MASLD (HR = 1.02, 95% CI = 0.88 – 1.19, p = 0.75) (**Fig 4B/C)**. We observed similar patterns of attenuation for adjustment for lifetime median HbA1c, and adjustment for treatment for T2D (defined as having been prescribed antidiabetic drugs on 6 or more occasions). Supporting this, we observed no association between r*s33950507* and cerebrovascular disease or MASLD in separate models restricted to individuals who did not receive a T2D diagnosis or treatment (**Fig 4C)**. The attenuation of the association between *rs33950507* and both CVD and MASLD outcomes in these sensitivity analyses therefore suggests the negative association may in part be mediated by HbA1c overestimation, T2D overdiagnosis, and over-treatment. Details of further sensitivity analyses are given in the **Supplementary Text.**

## Discussion

In this analysis of 43,088 British south Asian individuals with linked whole-exome sequencing and longitudinal electronic health record data, we show that genetically defined β-thalassaemia trait is prevalent but underdiagnosed in routine clinical care – 3 in 4 genetically-defined thalassaemia trait carriers did not have a clinical diagnosis. Thalassaemia trait influences HbA1c independently of blood glucose level and alters type 2 diabetes diagnosis and clinical outcomes: a common structural HBB variant (HbE) was associated with higher HbA1c, increased hazards of prediabetes and T2D diagnosis, but lower risk of several diabetes-related complications, likely mediated via over-diagnosis and over-treatment of T2D.

A growing body of evidence has described associations between genetically-determined erythrocytic traits and HbA1c values across genes including *G6PD, PIEZO1, and HBA1/2,* with a high impact on global populations due to ancestral variation. ^1,2,32^. To date, these genetic variants have been associated with a non-glycaemic reduction in HbA1c, which has been attributed to variation in red blood cell biology.

In this analysis, and to our knowledge for the first time, we describe a common pathogenic genetic variant in the *HBB* gene (causative of HbE trait), which is not associated with blood glucose, but is associated with increased HbA1c **(Fig 2)**, increased risk of prediabetes and T2D **(Fig 3A)**, and lower risk of T2D-related complications including cerebrovascular disease and eye disease **(Fig 4A)**. These findings are directionally opposite to reported associations between T2D-related outcomes and variants associated with an erythrocytic reduction in HbA1c. This provides further evidence for the hypothesis that disruption between glucose and HbA1c in red cell conditions can have meaningful effects on T2D diagnosis and progression. The combination of increased risk of prediabetes diagnosis, modestly increased hazards of T2D diagnosis, and reduced progression from T2D to diabetes-related complications is most parsimoniously explained by HbA1c-driven overclassification and overtreatment rather than increased underlying diabetogenic risk, as supported by sensitivity analyses following a mediation analysis framework.

It has been previously reported that HPLC methods of HbA1c measurement overestimate HbA1c in HbE^20^ due to changes in cationic charge causing assay interference. While HPLC-measured HbA1c may be falsely elevated in HbE trait carriers, other techniques such as immunoassay in these individuals may be at risk of the same disruption between red cell stability, glycaemia, and HbA1c values^15,33,34^. When we attempted to meta-analyse findings in UK Biobank, we discovered that HPLC-measured baseline HbA1cs for individuals with HbE trait were excluded from analysis with an “analyser result deemed not reportable” flag. This suggests that quality control for entry into research studies may be more rigorously applied than in clinical care, and offers insight into the need better quality assurance in routine diagnostic laboratories to avoid false diagnosis of prediabetes or T2D in HbE trait carriers. Overdiagnosis is well-reported to associated psychological harms^35^, excess healthcare costs^36^, overmedicalisation, and initiation of the testing cascade^37^, and is at odds with patient-centred care and minimally disruptive medicine aims^38,39^. These findings also have particular relevance to individuals who are older, or who have multiple comorbidities or frailty^40^, who are particularly at risk of harm from overtreatment^41^. However, it could also be argued the reduced risk of complications represented a beneficial element to T2D overdiagnosis and/or treatment.

For rare variants in *HBB*, analysed collectively in an aggregated mask to boost power due to low allele frequencies, we observe a similar pattern to that previously described in *G6PD* and *PIEZO1*^1,42^, with non-significant trends towards lower HbA1c **(Fig 2)**, and increased hazards of diabetic eye disease **(Fig 4A),** suggesting complexity and the possibility of mechanistically distinct pathways across the allele frequency spectrum.

In addition, our study highlights the low diagnosis rate (28.9%) of thalassaemia trait in a high risk south Asian population in the UK, who are not routinely screened for the condition outside of pregnancy, in keeping with previous global reports of underdiagnosis of thalassaemia trait^26^. Identifying these individuals without widespread genetic testing presents a challenge given an existing clinical risk prediction tool (ThalPred)^27^ did not effectively discriminate thalassaemia trait carriers from non-carriers in the context of microcytic, hypochromic anaemia in our study. These findings suggest that even laboratory-based discrimination strategies are insufficient to reliably identify carriers in this population, raising the possibility that genomic ascertainment or routine haemoglobinopathy screening in specific populations may be required to mitigate biomarker misclassification through a precision medicine approach^43^. Beyond implications for HbA1c interpretation, under-recognition of thalassaemia trait may also have broader clinical consequences, including misclassification of microcytic anaemia as iron deficiency, and missed opportunities for reproductive counselling. Although not the primary focus of this study, these findings underscore the wider clinical relevance of improved ascertainment.

Strengths of this study include its scale, focus on an underrepresented population, and linkage of cutting-edge exome sequencing with real-world data to explore genetic factors associated with real-world clinical outcomes. Its weaknesses include limited external replication beyond previously published summary statistics, and reliance on real-world electronic health record data for case and control ascertainment, with associated risks of ascertainment bias, survival bias, miscoding, and misclassification. Given the number of complication outcomes examined, nominally significant positive findings which do not survive correction for multiple testing should be interpreted cautiously and warrant replication in independent cohorts.

In summary, these findings suggest that for individuals with HbE beta-thalassaemia trait, reliance on HbA1c to diagnose, monitor and manage prediabetes and T2D may be associated with overdiagnosis and over-treatment, potentially affecting hundreds of millions of individuals on a global scale^7^. Because most individuals with genetically-defined thalassaemia trait are undiagnosed, many patients and clinicians will not be aware of the relevance of this to their care. In populations in which haemoglobinopathy variants are common, reliance on HbA1c, a biomarker of haemglobin glycation not direct measure of glycaemia may introduce systematic bias and inequity in diagnosis and care at scale. Alongside evidence from other erythrocytic conditions such as G6PD deficiency, these findings highlight the urgent need for enhanced quality assurance of HbA1c testing and alternative approaches to diagnosis and monitoring of T2D, particularly in ancestrally diverse populations, among whom erythrocytic traits are more common.

## Funding

SH is funded by Wellcome Health Advances in Underrepresented Populations (HARP) Doctoral Fellowship 227532/Z/23/Z. DS and SF are funded by the Tackling Multimorbidity at Scale Strategic Priorities Fund programme (MR/W014416/1) delivered by the Medical Research Council and the National Institute for Health Research in partnership with the Economic and Social Research Council and in collaboration with the Engineering and Physical Sciences Research Council. IB, TJM and SF are funded by a Wellcome Discovery Award (227897/Z/23/Z) and IB and TJM acknowledge support from the National Institute for Health and Care Research Exeter Biomedical Research Centre. RM is supported by Barts Charity (MGU0504). JT is supported by UKRI/Medical Research Council UKRII814. MKS is supported by Barts Charity grants MGU0504 and G-002995.

Genes & Health is/has recently been core-funded by Wellcome (WT102627, WT210561), the Medical Research Council (UK) (M009017, MR/X009777/1, MR/X009920/1), Higher Education Funding Council for England Catalyst, Barts Charity (845/1796), Health Data Research UK (for London substantive site), and research delivery support from the NHS National Institute for Health Research Clinical Research Network (North Thames). We acknowledge the support of the National Institute for Health and Care Research Barts Biomedical Research Centre (NIHR203330); a delivery partnership of Barts Health NHS Trust, Queen Mary University of London, St George’s University Hospitals NHS Foundation Trust and St George’s University of London Genes & Health is/has recently been funded by Alnylam Pharmaceuticals, Genomics PLC; and a Life Sciences Industry Consortium of AstraZeneca PLC, Bristol-Myers Squibb Company, GlaxoSmithKline Research and Development Limited, Maze Therapeutics Inc, Merck Sharp & Dohme LLC, Novo Nordisk A/S, Pfizer Inc, Takeda Development Centre Americas Inc.

The views expressed are those of the authors and not necessarily those of the NIHR or the Department of Health and Social Care, nor any other funder.

### Ethical Approval

A favourable ethical opinion for the main Genes & Health research study was granted by NRES Committee London - South East (reference 14/LO/1240) on 16 Sept 2014. Queen Mary University of London is the Sponsor, and Data Controller.

## Supporting information

Supplementary Methods & Results

## Acknowledgements

For the purpose of open access, the author has applied a Creative Commons Attribution (CC BY) licence to any Author Accepted Manuscript version arising from this submission.

We thank Social Action for Health, Centre of The Cell, members of our Community Advisory Group, and staff who have recruited and collected data from volunteers. We thank the NIHR National Biosample Centre (UK Biocentre), the Social Genetic & Developmental Psychiatry Centre (King’s College London), Wellcome Sanger Institute, and Broad Institute for sample processing, genotyping, sequencing and variant annotation. This work uses data provided by patients and collected by the NHS as part of their care and support. This research utilised Queen Mary University of London’s Apocrita HPC facility, supported by QMUL Research-IT, http://doi.org/10.5281/zenodo.438045

We thank: Barts Health NHS Trust, NHS Clinical Commissioning Groups (City and Hackney, Waltham Forest, Tower Hamlets, Newham, Redbridge, Havering, Barking and Dagenham), East London NHS Foundation Trust, Bradford Teaching Hospitals NHS Foundation Trust, Public Health England (especially David Wyllie), Discovery Data Service/Endeavour Health Charitable Trust (especially David Stables), Voror Health Technologies Ltd (especially Sophie Don), NHS England (for what was NHS Digital) - for GDPR-compliant data sharing backed by individual written informed consent.

## Most of all we thank all of the volunteers participating in Genes & Health

### Data availability

Genes & Health: Individual-level participant data are available to researchers and industry partners worldwide via application to and review by the Genes & Health Executive (https://www.genesandhealth.org/); applications are reviewed monthly. Approved researchers have access to individual-level data in the Genes & Health Trusted Research Environment (TRE) and can request the data files used in this study from the corresponding author(s). All data exports from the Genes & Health TRE are reviewed to prevent release of identifiable individual-level data. Summary data may be exported for cross-cohort meta-analysis or replication and for publication, subject to review.

