## Supplementary Methods & Results for "HbA1c-based diagnosis of type 2 diabetes and complication risk are distorted in British south Asians due to HbE thalassaemia trait"

**Thalassemia Paper Supplementary Material**

### Supplementary Methods

#### Whole exome sequencing

In brief, sequencing was performed using the Twist Biosciences platform with Illumina 150bp PE Novaseq-6000 sequencing; crams were produced after mapping to hg38, and only those with coverage > 85% at >20x were included. Pakistani and Bangladeshi ancestry were defined using UMAP and correlated highly with population references from 1000 Genomes; individuals not corresponding to these ancestral groups were excluded (n = 198). For variant-level QC a random forest model was trained on a set of variants corresponding to TOPMED-imputed genotype data from the same individuals (ie, true positive concordance). Further details are provided in[1].

#### Complication definitions

We defined microvascular complications of type 2 diabetes as chronic kidney disease, diabetic neuropathy, or diabetic eye disease. Microvascular complications were defined as coronary artery disease, heart failure, peripheral arterial disease, or cerebrovascular disease. Because of high prevalence and related pathophysiology of metabolic-dysfunction associated steatotic liver disease (MASLD) in our cohort, we additionally examined association of thalassemia genotypes with this outcome.

For individuals with a diagnosis of T2D, complications of T2D were defined only as those complications defined above which first appeared in the electronic health record on a date after the earliest date of type 2 diabetes diagnosis. For sensitivity analyses in individuals free from T2D (see below) we defined complications using the same codelists, but not predicated upon a pre-existing T2D diagnosis.

#### Antidiabetic medication

We defined antidiabetic medication as prescriptions in the primary care record from the British national Formulary (Chapter 6, Section 6.1: Drugs used in diabetes)(**Table S4)**. We selected the earliest date of prescription of any antidiabetic medication as the “earliest date” before which pre-diabetic HbA1c and glucose values must be obtained on, accepting that some non-diabetic HbA1c and glucose readings would be lost by this conservative approach in, for example, patients with transient gestational diabetes, polycystic ovarian syndrome, or prescribed drugs such as SGLT2 inhibitors for alternative indications such as heart failure.

In sensitivity analyses, we combined all antidiabetic medication from Chapter 6, Section 6.1 to create a binarized “ever/never” phenotype of “having been prescribed antidiabetic drugs on 6 or more instances, as a reasonable proxy for exposure to diabetes treatment. In sensitivity analyses, the models we ran were unchanged when the number of prescriptions was varied between 1-12.

#### HbA1c assays

For HbA1c values included in this study, recorded values largely represent our participants based in north East London where HbA1c is analysed using high-performance liquid chromatography (HPLC) from a Biorad HbA1c D100 analyser at the NHS East and South-East London Pathology Partnership with accreditation from the UK Accreditation Service (reference 8285) However, it is also possible that small numbers of included HbA1c values reflect analysis at other sites (eg by patients moving between geographic areas, or entered from the national diabetes audit); however, the exact number of these is not identifiable as the origin of each HbA1c is not recorded within the routine datasets used in this analysis.

Association testing: single variants with minor allele frequency >1%

For common genetic variants with MAF >1%, we described the association of carrier status with quantitative traits using multivariable logistic regression models adjusted for age at study recruitment, age at test (for clinically measured quantitative variables), sex, genetically-defined ancestry, and the first 20 genetic principal components. For similarly-adjusted multivariable survival analyses, we described the association of carrier status with progression from birth to pre-diabetes and type 2 diabetes; and progression from type 2 diabetes diagnosis to diabetes-related complications. Individuals were censored at the earliest of date of death, date of last data extraction, or the outcome in question. We additionally explored univariate associations using Kaplan-Meier plots. Both multivariable logistic regression and survival analyses were run using REGENIE V4.0 with the "—t2e" flag under an additive model, to allow direct comparison with gene-based results, which could not be performed using traditional cox proportional hazards models in other software.

Gene-based association testing

For rare variants with a MAF <1%, the relatively low count of affected individuals for each variant precludes association testing as it predisposes to case-control imbalance. This can be overcome by "collapsing" all rare variants with similar annotations lying within a gene into a single aggregated group, commonly called a "mask". The association of this mask with outcomes of interest can then be explored using REGENIE v4.0 using multivariable regression and survival models, adjusted as described above.

Meta-Analysis of results with publicly available summary statistics

For rare ClinVar pathogenic variants in *HBB* with minor allele frequency < 0.01 (HBB Rare LP/P), results were meta-analysed with those described from the most closely aligned mask available on the AstraZeneca phewas portal[2], composed of UK Biobank summary statistics (UKBB-ptv: protein-truncating variants). For the single HbE trait variant (rs33950507), we attempted to meta-analyse findings in UK Biobank. However, we discovered that HPLC-measured baseline HbA1cs for individuals with HbE trait were excluded from analysis with an "analyser result deemed not reportable" flag. Results for HbE trait were meta-analysed with whole exome sequenced results published as part of the AMP-T2D study[3].

Sensitivity analyses

In addition to the sensitivity analyses described in the main text, we explored the association of T2D complications ~ *HbE* genotype in individuals without a diagnosis of T2D in their EHR. We hypothesised that among these individuals the absence of the plausible mechanisms above should ameliorate the association between *HbE* trait and outcomes.

We attempted to exclude the possibility of other mechanisms in explaining the relationship between T2D complications and HbE genotype. First, we hypothesised that the increased risk of heart failure (HF) associated with HbE trait may act as a competing risk. Although we did not analyse mortality in the absence of high-quality data linkage, it is possible the increased HF risk may in turn associate with lower risk of other complications if, for example, individuals develop HF and die before they develop other complications. We undertook a competing risks survival analysis using the R package ]cmprsk[4] to explore this.

Finally, we hypothesised that different T2D complication rates in *HbE* trait may reflect differential diagnosis and interventions not explained by, for example, HbA1c. To test this, we explored the association of *HbE* genotype with (1) odds of type 2 diabetes diagnosis in individuals with 2 or more HbA1c >=48, and (2) time from T2D diagnosis to first receipt of antidiabetic medication, reasoning that similar effect sizes between HbE heterozygotes and non-thalassemia carriers would suggest different complication rates were not explained by differential exposure to healthcare diagnostics and intervention.

Finally, we hypothesised that different T2D complication rates in *HbE* trait may reflect differential diagnosis and interventions not explained by, for example, HbA1c. To test this, we explored the association of *HbE* genotype with (1) odds of type 2 diabetes diagnosis in individuals with 2 or more HbA1c >=48, and (2) time from T2D diagnosis to first receipt of antidiabetic medication, reasoning that similar effect sizes between HbE heterozygotes and non-thalassemia carriers would suggest different complication rates were not explained by differential exposure to healthcare diagnostics and intervention.

**Supplementary Results**

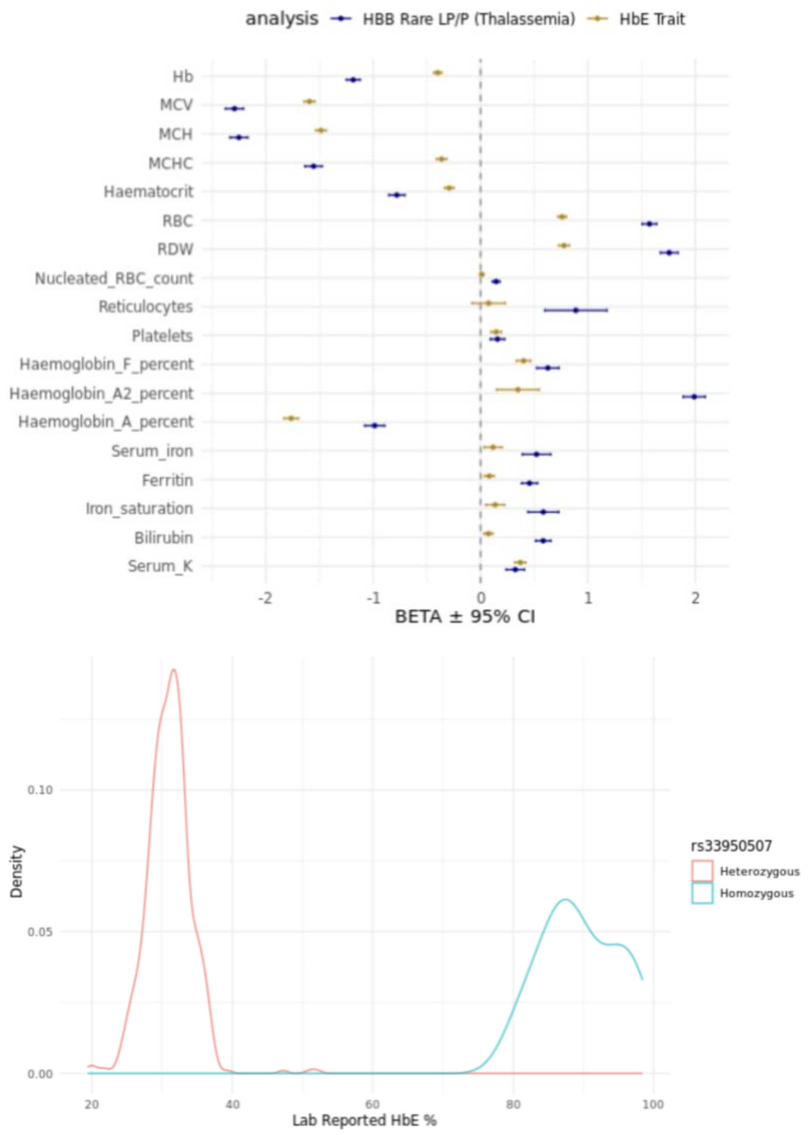


***<u>Figure S1: Association of pathological thalassemia HBB variants with red cell and related traits in 43,088 individuals with measured full blood count in the Genes & Health study. A:</u>*** *<u>Common and rare pathologic thalassemia variants in HBB are associated with directionally consistent erythrocytic changes. Associations are presented for a single common variant (rs33950507, causal for HbE trait) with MAF >1%, which is causal for HbE trait. 38 Clinvar pathogenic / likely pathogenic variants with MAF <1% were collapsed in to a single mask (HBB rare LP/P). B: Lab-reported HbE % values for 590 rs33950507 heterozygotes and 15 homozygotes, presented as density plots stratified by genotype. HbE% was available for 606 participants in total in the study; only 1 participant with a measured HbE% did not have at least one copy of the rs33950507 effect allele.</u>*

*HbE Trait is associated with reduced hazards of progression from prediabetes to T2D*

Because we observed a stronger relationship between HbE trait and prediabetes than HbE trait and type 2 diabetes (in terms of both greater magnitude of hazard ratio and smaller p value), we performed additional exploratory analyses investigating the relationship between HbE and transition from prediabetes to T2D. First, we observed no association between HbE trait and type 2 diabetes polygenic risk score (PRS)[5] in uni- or multivariate regression analyses, nor in ANOVA comparing HbE trait frequency across PRS deciles (**Fig S2).** HbE trait was, however, associated with reduced hazards of T2D among individuals diagnosed with prediabetes in multivariate survival models (HR = 0.60, 95% CI – 0.48 – 0.73, p = 2.42 x $10^{-7}$) and univariate analysis (**Fig S2).** In a separate multivariate logistic regression model restricted to individuals with prediabetes who subsequently developed type 2 diabetes, we observed no difference in time from prediabetes to type 2 diabetes between HbE trait carriers and unaffected individuals (beta = 0.56, 95% CI = -0.31 – 1.43, p = 0.56). Taken together, these findings suggest HbE trait is not associated with risk of type 2 diabetes per se, but could suggest that individuals diagnosed with prediabetes on the basis of HbE-associated HbA1c elevations may be at lower risk of progressing to type 2 diabetes than those without HbE. This could be because those individuals without HbE trait have underlying genetic or environmental risk factors which propose them to progressing on to type 2 diabetes. Alternatively, it could be that the combination of increased hazards of prediabetes diagnosis, modestly increased hazards of T2D diagnosis, and substantially reduced progression from prediabetes to T2D is most parsimoniously explained by HbA1c-driven overclassification rather than increased underlying diabetogenic risk.

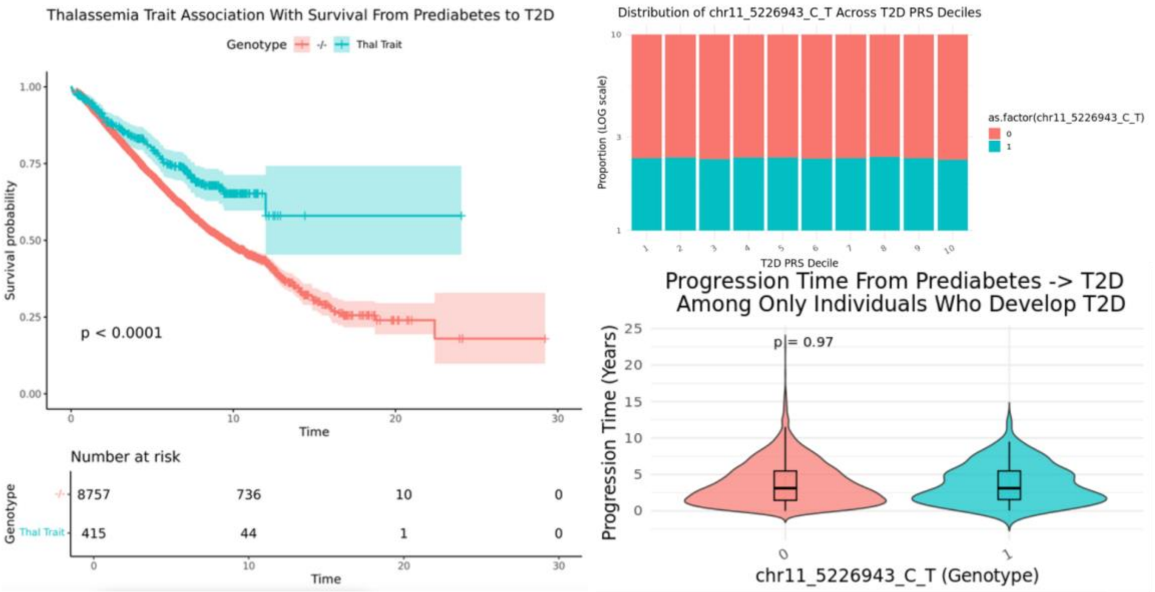


**Supplementary Figure 2: A: Kaplan-Meier plot depicting survival from date of prediabetes diagnosis to type 2 diabetes diagnosis, stratified by HbE genotype – HbE trait carriers (blue line) vs unaffected individuals (red line). B: Distribution of HbE genotype across 43,092 individuals included in analysis, stratified by type 2 diabetes polygenic risk score (T2D PRS) decile. C: Distribution of time from date of prediabetes diagnosis to date of T2D diagnosis among only those individuals from panel A who subsequently developed T2D, stratified by HbE genotype.**

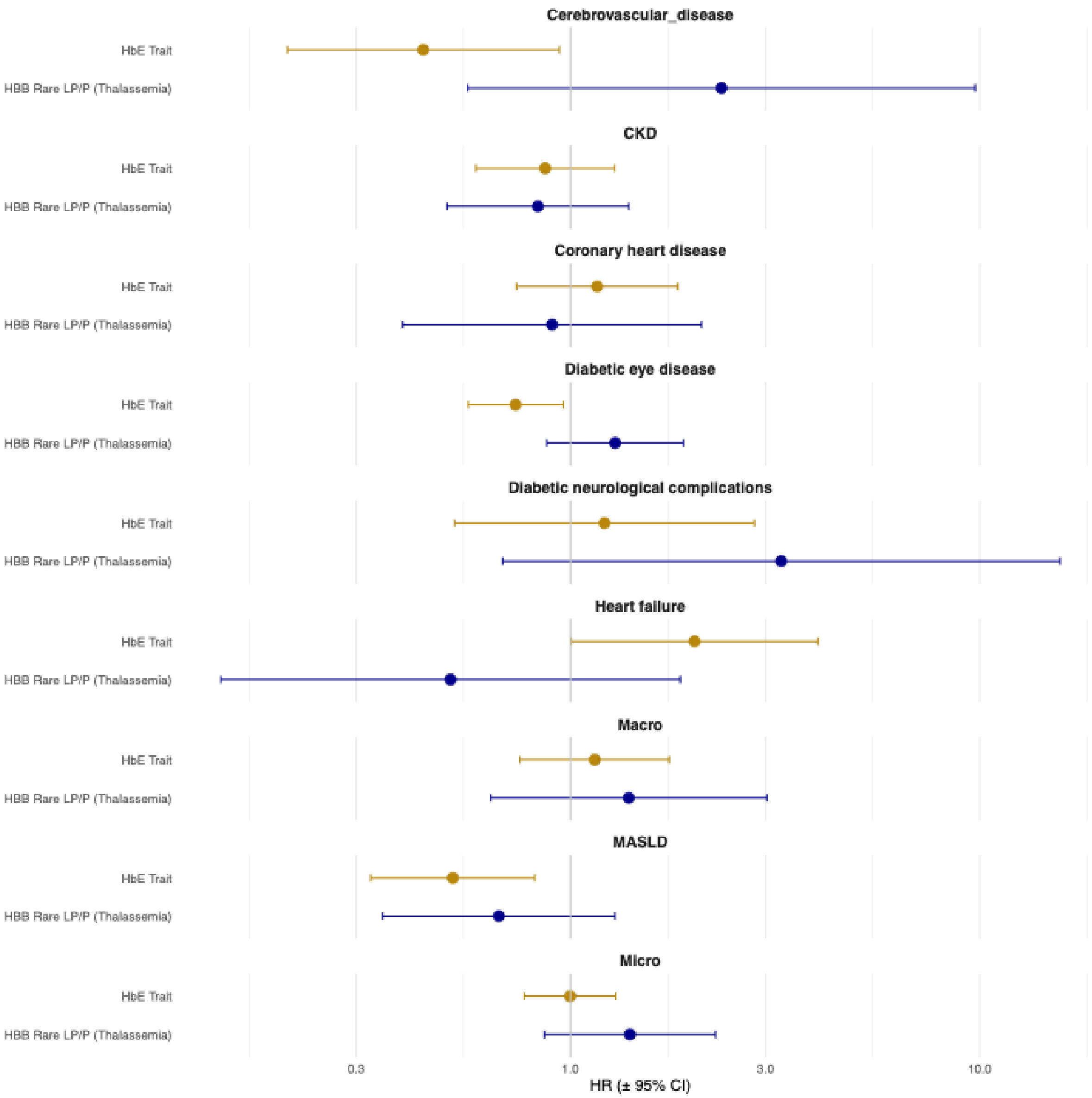


**Fig S3. Association of pathogenic thalassemia trait variants with progression from birth to diabetes-related complications. Results are presented as hazard ratio (HR) +- 95% confidence interval, estimated from multivariable Cox proportional hazards models adjusted for age at recruitment, age at recruitment squared, sex, genetically inferred ancestry, and the first 20 genetic principal components, presented separately for rare Clinvar likely pathogenic / pathogenic variants in *HBB* with minor allele frequency < 0.01 (HBB Rare LP/P), and a single common variant (rs33950507, causal for HbE trait). MASLD: metabolic dysfunction-associated steatotic liver disease.**

### Sensitivity analyses

In addition the sensitivity analyses reported in the main text, we did not observe any attenuation of CVD or MASLD associations in a competing risks survival model assessing the impact of (reduced) heart failure survival on these outcomes in HbE carriers (**Fig S3)**. Because differences in rates of diagnosis or treatment of T2D among individuals with elevated HbA1c could vary between r*s33950507* heterozygotes and non-carriers, and impact complication rates, we additionally explored whether differences in these variables could plausibly underlie the identified associations between rs33950507 and diabetes-related complications (**Fig S3)**. We identified no association between *rs33950507* and any of (1) survival to T2D diagnosis, (2) exposure to diabetes-controlling medication, or (3) survival from T2D diagnosis to medication initiation **(Fig S3).** Furthermore, we identified no association between rs33950507 and acne vulgaris, included as a negative control.

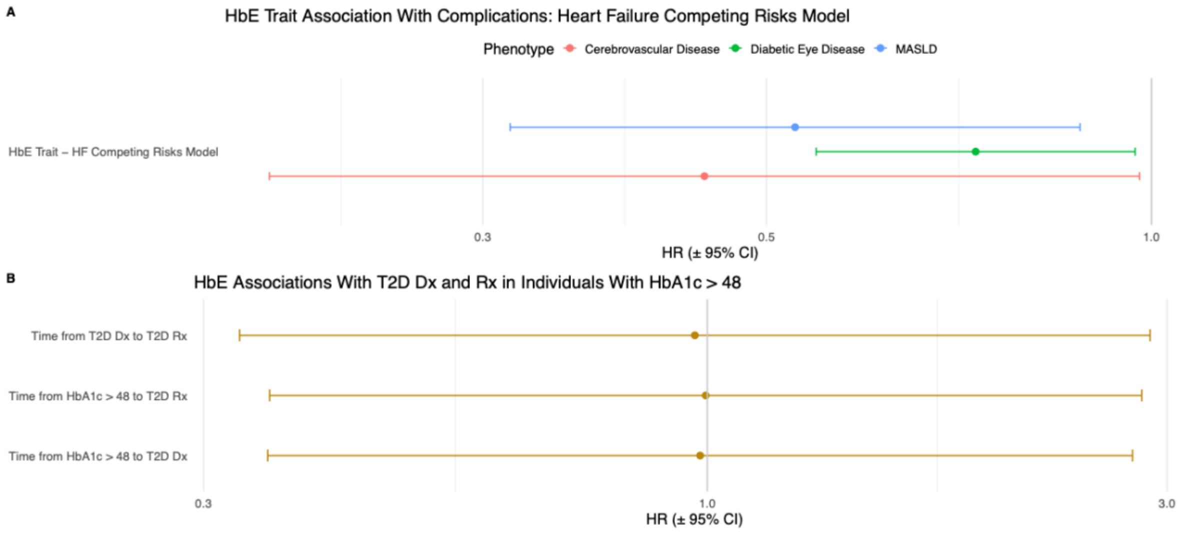


**Fig S4. Results from sensitivity analyses based on the analyses in Fig 3A describing associations between HbE trait and diabetes-related complications. A: competing risks model describing association of HbE trait with diabetes-related complications in a competing risks survival model describing the competing risk of heart failure diagnosis. B: Association of HbE trait with time from high HbA1c (>48mmol/mol) reading in the EHR to T2D Dx, initiation of T2D Rx, and time from T2D Dx to initiation of Rx.**
